# Self-reported reactions to the front-of-package warning labelling in Mexico among parents of school-aged children

**DOI:** 10.1101/2023.07.26.23293213

**Authors:** Carolina Batis, Tania C. Aburto, Lilia S. Pedraza, Erick Angulo, Zugey Hernández, Alejandra Jáuregui, Lindsey Smith Taillie, Juan A. Rivera, Simón Barquera

## Abstract

**Background:** To improve the food environment and guide consumers to select healthier foods, the implementation of a front of package warning labelling (FOPWL) started in Mexico in October 2020. We aimed to identify the self-reported support, understanding, use and perceived impact of the FOPWL 1-5 months after its implementation among parents of school-aged children across socioeconomic categories and nutrition knowledge and attitudes.

**Methods:** EPHA-niñ@s is a national web-based cohort of Mexican children 5-10 y and one of their parents aiming to monitor their food and food policy perception and opinions and children’s dietary intake. Recruitment was conducted primarily through paid advertisements on social media. Data was collected online with a self-administered questionnaire answered by the parent and an interviewer-administered questionnaire answered by the child during a video call. This analysis was conducted with data from the parent’s questionnaire from the first wave of data collection (November 2020-March 2021) which included 2,071 participants from all over the country. We evaluated differences by socioeconomic status (SES), education and nutrition knowledge and consciousness, while adjusting by other sociodemographic characteristics using multinomial logistic regression.

**Results:** The sample was predominantly from middle and high socioeconomic status (SES). Most parents (85%) agree/strongly agree with the FOPWL (support), 86% correctly identified that a product with one warning is healthier than a product with three (understanding), 65% compared the number of warnings sometimes to very often (use), and 63% reported buying less and 25% stopped buying products with warnings for their children (perceived impact). The perceived impact was higher when products were for their children than for themselves. Perceived impact also differed by food group, being higher for sodas, juices, and cereal bars and lower for chips and chocolate powder. Responses were more favorable for five-six questions (out of seven) among those with higher nutrition knowledge, and higher nutrition consciousness, and for three questions among those with higher education level.

**Conclusion:** Within six months of implementation, the immediate self-reported responses related to support, understanding, use, and perceived impact to the Mexican FOPWL were favorable. Further studies in other populations including low SES participants and impact evaluations, are needed.

## BACKGROUND

The health and economic burdens due to childhood obesity and poor dietary habits among the youth are a key pressing matter in Mexico and elsewhere. The prevalence of overweight and obesity in school-aged children in Mexico is 32%, the 6^th^ largest in OECD countries [1,2]. Mexico is the country with the 4th highest per capita purchases of ultra-processed snacks, breakfast cereals, and sugar-sweetened beverages [3]. The latter results are particularly worrisome among children and adolescents, whose caloric intake from ultra-processed foods reach34—38% of their total daily energy intake (in adults is 26%) [4] The excessive intake of unhealthy foods and beverages is related to the unregulated and aggressive food environment generated by transnational food corporations [5–7]. This unhealthy food environment cannot be counteracted effectively by the population that lacks the resources, such as nutrition education, to eat healthy [8]. Meanwhile, the health and economic loss due to obesity are severe, as obesity reduces life expectancy by 4.2 years and the country’s GDP by 5.3%[9].

To improve the food environment and guide consumers to select healthier foods, a new front-of-package labelling was recently implemented in Mexico [10]. Several front-of-package labelling models have been proposed, such as graded star systems, color scales, letter grades, traffic lights, or stop signs-type warning labels [11]. Research suggests that front-of-package warning labelling (FOPWL) such as the one first introduced in Chile, is likely the most effective [12]. In the Mexican population of different socioeconomic backgrounds, several experiments and qualitative studies were conducted to test different FOP labelling models [scores, traffic lights, warning labels (high-in)][13–15]. Consistently, it was found that the FOPWL was amongst the models most easily understood by the population and most effective in improving the healthfulness of purchases within experimental settings. The FOPWL implemented in Mexico in October 2020 comprises octagon-shaped black warnings displaying “excess-in” for five nutrients (calories, added sugars, saturated fat, trans fat, and sodium). Nutrient thresholds are mainly based on the ones proposed by the Pan American Health Organization (PAHO)[16]; and will become stricter in Phase 2 (October 2023) and 3 (October 2025) of implementation. It also includes cautionary legends for products containing caffeine or artificial sweeteners (Figure 1) advising to be avoided by children. Since mid-2021, products with one or more warnings or cautionary legends cannot display cartoon characters or health claims. So far, the FOPWL has not been accompanied by the implementation or strengthening of any other policy or a national communication campaign [10].

**Figure 1.**
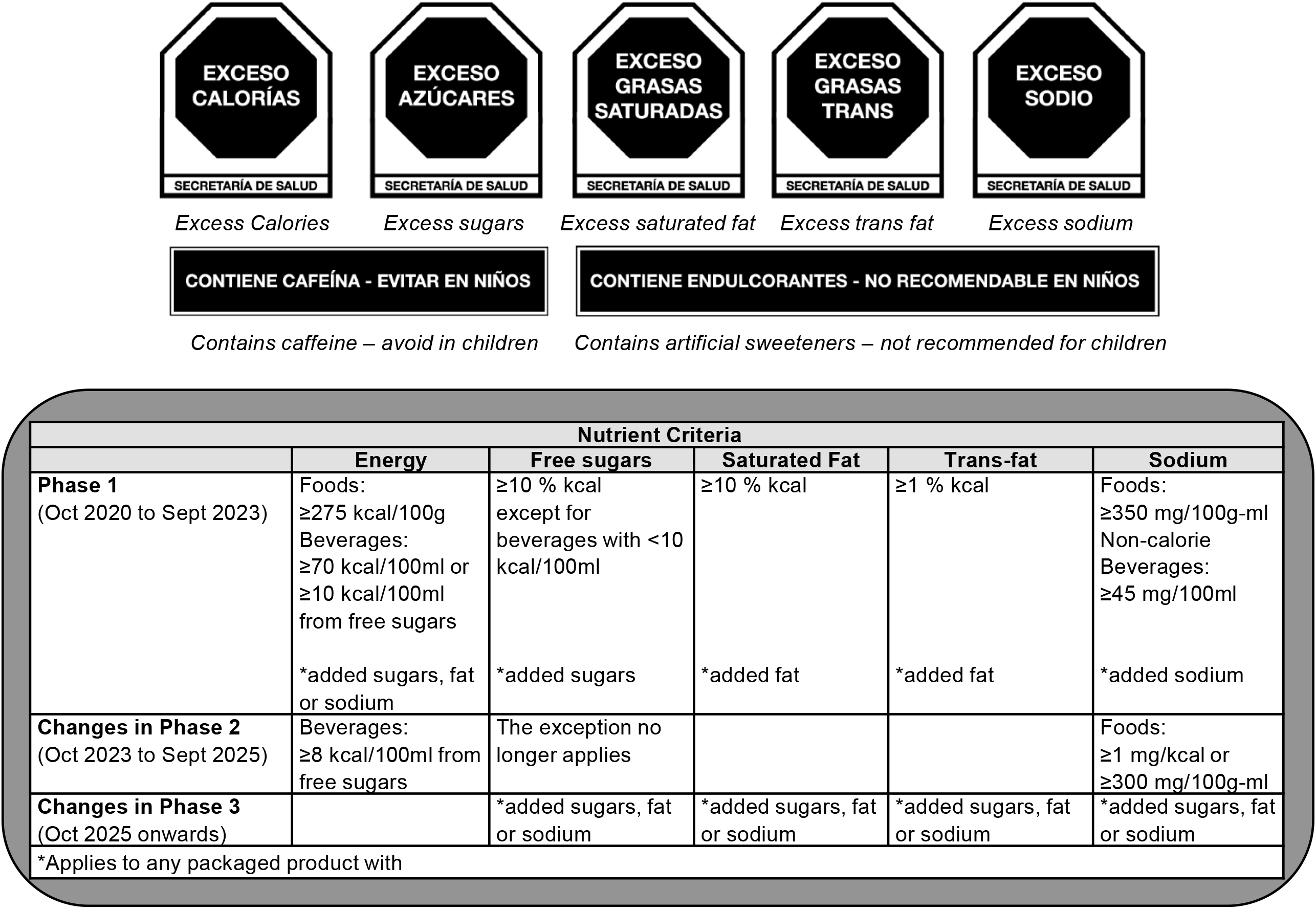
Summary of Mexican FOPWL.

FOPWL is considered an effective public health tool to prevent obesity, both by modifying individual’s food choices and by motivating the food industry to reformulate their products [17,18]. Warnings are meant to rapidly inform the consumer about nutritional characteristics of the product (e.g., “high in sugar/sodium/saturated fat” signs) and discourage the purchase and intake of unhealthy products [19]. To work, the FOPWL should first be noticed and understood, then it should elicit negative affect, increase perception of risk, trigger behavioral intentions, and ultimately it should provoke behavioral change [20]. Nonetheless, FOPWL is an agento-structural intervention, meaning that the structural change produced by the intervention facilitates healthier choices but still relies on individuals’ decisions [21]. Preexisting food choices as well as food and health values, attitudes towards food, nutrition knowledge, education level and socioeconomic status (SES) are all factors that can moderate how effective FOPWL is [20]. Therefore, it is important to evaluate if the response to the FOPWL differs by individual characteristics. Furthermore, evaluating the FOPWL among parents is of particular interest given their key role in children’s dietary intake. The parents experience with the FOPWL, and the influence that this has on their purchasing decisions is unique, as it includes considerations related to the health and the likings of themselves and their children[22,23].

Mexico is joining a shortlist of countries (Chile, Peru, Uruguay, and Israel) that have FOPWL; hence empirical evidence is just emerging. In Chile, FOPWL along with marketing and school regulations were implemented since 2016 [24]. Studies from Chile found that the FOPWL was accepted by the population, that it changed their knowledge and perception of foods, and that the purchases of ‘High-in’ beverages decreased 24% and the content of sugar, saturated fat, and sodium from the overall purchases decreased 10, 4 and 5%, respectively [25–28]. Given the limited real-life evidence worldwide and the unique characteristics and context of the Mexican FOPWL, evaluations of this policy are required. Studies evaluating intermediate outcomes (e.g., understanding, behavioral intentions, and reformulation) are useful, particularly because final outcomes (food purchases, diet quality, or obesity) are hard to evaluate for national policies that lack a control group [17]. Therefore, it is key to document the population’s response to the new FOPWL in Mexico, particularly among those involved in shaping the children’s dietary habits. Hence, we conducted this study among parents of school-aged children participating in a national web-based cohort. The aim of this study was to identify the support, use, perceived impact, and understanding of the new FOPWL among all parents participating in the study and across sociodemographic variables and nutrition consciousness and knowledge.

## METHODS

### Study population

The study population is from the EPHA-niñ@s cohort (Study of the Perception and Dietary Habits in Children, for its acronym in Spanish), a recently established online cohort of Mexican children aged 5-10 y old and one of their parents or primary caregiver (hereafter referred as “parents”). Participants are distributed across the 32 states of Mexico. The overall purpose of EPHA-niñ@s is to monitor parents’ and children’s food perceptions and preferences and children’s dietary intake over time, with the goal of evaluating national-level food policies such as FOPWL, school guidelines and health education, marketing, sales regulation, and taxes. Data for this analysis comes from the first wave of data collection of EPHA-niñ@s. The first wave was conducted from November 2020 to March 2021, right after the implementation of the FOPWL in Mexico, and its questionnaire was mainly focused on specific FOPWL intermediate outcomes (i.e., support, use, perceived impact, and understanding).

Parent-child dyads were eligible if they both lived in Mexico and had no plans of changing residence abroad in the following years, had internet access at home (not only from cellphone), the child was between 5 to 10 y old and did not have any major chronic conditions, eating disorders, or food allergies, and parents were not currently working for the food industry. Study participants were recruited through paid advertisements from the social media accounts of the study. As a reference, in Mexico 74% of adults aged 25-34 years and 57% of adults aged 35-44 years are social media users[29]. To enhance the reach, the study posts were also shared through UNICEF’s social media accounts, and information about the study was given in one of the Ministry of Education webinars to parents and teachers nationwide. Information about the study was not disseminated through our institutional social media accounts (INSP, National Institute of Public Health, or CINyS, Health and Nutrition Research Center), as the followers of these accounts might be highly aware or educated on nutrition topics and food policies (the type of content shared in these accounts). In Mexico,

The study protocol was reviewed and approved by the Research and Ethics Committees of the INSP. The study advertisements had a link to the inclusion criteria questionnaire and if eligible, a link to the inform consent form. Subjects agreeing to participate were then given access to the full online self-administered questionnaire (∼40 minutes). A personalized link of their questionnaire was sent to their email, so that participants could be able to save and continue with the questionnaire during several sessions. Participants were also contacted by cellphone and invited to schedule a video call with their participating child and a research staff member. During the video call an interviewer-administered questionnaire was conducted children’s food perception and preferences. In the first wave of data collection, 6895 participants completed the inclusion criteria and were eligible, 4036 signed the informed consent, 2182 completed the online questionnaire and 1872 had a video call. For this analysis, we included all participants that completed the online questionnaire and had valid information on the variables of interest (n=2071).

### Measurements

All measurements included in this analysis were collected through the self-administered questionnaire answered by the parent. All questions were multiple-choice and close-ended. The questions of interest in this analysis were related to the FOPWL: 1*) Support* was captured with a question about whether they agreed with the FOPWL (response options ranged from strongly agree to strongly disagree). 2) *Understanding* was captured with three questions, two captured subjective understanding (i.e., how easy/hard to understand they thought the FOPWL was for themselves and for their children), while the third one measured objective understanding by asking parents to choose the healthier option between a product with three warnings and a product with one warning. Additionally, an item queried whether they had seen or heard on the media information about the FOPWL. 3) *Use* was measured with a question about how often they compared the number of warnings between similar products (response options ranged from never to very often). 4) *Perceived impact* was assessed with three sets of questions. The first set included two questions inquiring about what parents have done when a product they used to buy had warnings, one question asked about the products they used to buy for their child and another about the products they used to buy for themselves (response options: have continued to buy it as before, have bought less than before, have stopped buying it, and have not seen warnings on the products I buy). The second set listed 13 food categories that frequently have warnings (sodas, bottled juices, chocolate powder, flavored milk, breakfast cereal, cookies, cereal bars, packaged peanuts, chips, yogurt, packaged cheese, processed meats, and packaged corn “tostadas”), and asked if they used to purchase each category regularly before the FOPWL. If affirmative, they were asked if the FOPWL made them change their decision to buy the products (response options: yes, stop purchasing it; yes, purchase less; yes, switch to products with less warnings; yes, switch to products with same warnings but different price; no, purchase the same because it has few/none warnings; and no, purchase the same because it is my favorite brand). The third set inquired whether a particular nutrient warning (calories, added sugars, saturated fat, trans fat or sodium), none, or all of them had influenced their purchase decisions.

As covariates we included geographical region, children’s age and sex, and parent’s sex, age, education level, and marital status. Parents were also asked about their weight and height with an “I don’t know” option. BMI was estimated and categorized as less than 25 kg/m^2^, overweight, obesity and weight/height not reported. The household SES was assessed using the Mexican Association of Market Research Agencies and Public Opinion Index [30]. This index classifies households into seven strata (from higher to lower: A/B, C+, C, C-, D+, D, E) based on six variables (number of bathrooms, bedrooms, vehicles, household members working, internet connection, and head of household education level). Using this index allowed us to compare the SES of our sample to that of the general Mexican population. For a reference, in the A/B category all have at least one car and full bathroom (70-80% have two or more from each), in the C the majority have only one from each, and in the E the majority have none. Additionally, the importance attributed to health and nutrition was collected by asking parents how often they choose their children’s foods according to their healthfulness. This variable was classified as high health nutrition consciousness: always/almost always; medium: sometimes; low: rarely/never. Finally, to capture nutrition knowledge, we asked parents their healthfulness perception for a set of 11 foods, including six healthy foods (mango, carrot, banana, zucchini, beans, and corn tortilla) and five less healthy foods (flour tortilla, turkey frankfurter, cereal bar, chocolate milk, and cured ham). For this purpose, parents were asked whether they agreed if each food was healthy, with five response options ranging from strongly agree to strongly disagree. Based on the food and option selected, an overall nutrition knowledge score was estimated. For healthy foods, 5 points were given if “strongly agree” was selected, 2.5 for “agree”, 0 for “nor agree/nor disagree”, −2.5 for “disagree”, and −5 for “strongly disagree”. For less healthy foods the points were reversed (−5 “strongly agree” to 5 “strongly disagree”). Total points were added, and the sample was divided into tertiles (high, medium, and low nutrition knowledge).

### Statistical analysis

Descriptive statistics were calculated to show the distribution of sample’s characteristics. For the variables related to the support, understanding, use, and perceived impact of FOPWL, we estimated the proportion of participants reporting each answer, we collapsed some answers into a single category (e.g., strongly agree and agree) to ease interpretation. We evaluated whether the responses differed by SES, education level, nutrition consciousness, and nutrition knowledge, adjusting by all other samplés characteristic. For this, we ran multinomial regression models, with the response to the FOPWL-related question as the dependent variable, and education level, socioeconomic status, nutrition consciousness, nutrition knowledge and other sample’s characteristics such as age and gender (of the child and parent), marital status, geographic region, and parent’s BMI as the independent variables. From these models we obtained predicted probabilities and chi-squares to test if the proportions differed by education level, socioeconomic status, nutrition consciousness and nutrition knowledge. For all analyses, we used a p-value <0.05 to consider results statistically significant. The analysis was conducted in STATA 15 (StataCorp, College Station, TX).

## RESULTS

### Sample characteristics

A third of the sample had a high SES (A/B: 33%) and most (58%) of the remaining sample had a medium SES (C+ to C-) (Table 1). Thirty-seven percent of the parents had college or higher education level, 76% were married, and 92% were females. There were participants from all regions of the country, and the age and sex of the children were balanced. A third of the parents did not report their weight/height, and from those reporting it, 62% were classified with overweight or obesity. Most of the parents (70%) self-reported being nutrition conscious (i.e., they almost always or always choose their children’s foods according to their healthfulness).

**Table 1.** Parent’s characteristics (n= 2,071) of the EPHA-niñ@s cohort (Study of the Perception and Dietary Habits in Children, for its acronym in Spanish).

|  | n | % |
| --- | --- | --- |
| Children's age |  |  |
| 5-6 y | 713 | 34.4 |
| 7-8 y | 751 | 36.3 |
| 9-10 y | 607 | 29.3 |
| Children's gender |  |  |
| Female | 1,070 | 51.7 |
| Male | 1,001 | 48.3 |
| Parents' age |  |  |
| <30 y | 577 | 27.9 |
| 30-39 y | 1,040 | 50.2 |
| ≥40 y | 454 | 21.9 |
| Parents' gender |  |  |
| Female | 1,899 | 91.7 |
| Male | 172 | 8.3 |
| Socioeconomic status (SES) |  |  |
| A/B (higher) | 689 | 33.3 |
| C+ | 684 | 33.0 |
| C | 518 | 25.0 |
| C- to E (lower) | 180 | 8.7 |
| Parents' education |  |  |
| Less than high school | 637 | 30.8 |
| High school (complete/incomplete) | 641 | 31.0 |
| Greater than or equal to college degree | 762 | 36.8 |
| Marital status |  |  |
| Married | 1,563 | 75.5 |
| Not married | 508 | 24.5 |
| Geographic area |  |  |
| North | 437 | 21.1 |
| Center | 704 | 34.0 |
| Mexico City | 221 | 10.7 |
| South | 709 | 34.2 |
| Parents' BMI |  |  |
| Underweight/Normal weight | 530 | 25.6 |
| Overweight | 562 | 27.1 |
| Obese | 285 | 13.8 |
| Don't know/didn't answer | 694 | 33.5 |
| Health nutrition consciousness |  |  |
| Never/rarely | 195 | 9.4 |
| Sometimes | 451 | 21.8 |
| Almost always/always | 1,425 | 68.8 |

### Support

Most parents (85%) strongly agree/agree with the FOPWL implementation (Table 2). Those with higher education and lower SES were less neutral, meaning a higher proportion both agreed and disagreed with the FOPWL in comparison to those with lower education or higher SES.

**Table 2.** Support, understanding, use, and perceived impact and to FOPWL reported by parents, among all and by sociodemographic characteristics*.

| Table 2: Support, understanding, use, and perceived impact and to FOPWL reported by parents, among an and by sociodemographic characteristics |  |  |  |  |  |  |  |  |  |  |  |  |  |  |
| --- | --- | --- | --- | --- | --- | --- | --- | --- | --- | --- | --- | --- | --- | --- |
|  | All | Caregiver education level |  |  | Socioeconomic status (SES) |  |  |  | Nutrition consciousness |  |  | Nutrition knowledge |  |  |
|  |  | <High school | High school | ≥College | C- a E (low) | C | C+ | A/B (high) | Low | Med | High | Low | Med | High |
| **Support** |  |  |  |  |  |  |  |  |  |  |  |  |  |  |
| Do you agree with FOPWL? |  | **p= 0.0093** |  |  | **p= 0.0253** |  |  |  | **p= 0.2461** |  |  | **p= 0.6206** |  |  |
| Strongly agree/agree | 85.1 | 84.9 | 84.3 | 88.2 | 88.1 | 87.1 | 86.7 | 84.1 | 86.0 | 83.3 | 87.3 | 85.8 | 85.3 | 88.4 |
| Neutral | 12.9 | 13.9 | 14.9 | 9.1 | 6.7 | 11.4 | 11.8 | 15.0 | 12.7 | 15.4 | 11.3 | 12.8 | 13.1 | 10.4 |
| Disagree/strongly disagree | 2.0 | 1.1 | 0.8 | 2.7 | 5.2 | 1.5 | 1.4 | 1.0 | 1.4 | 1.3 | 1.5 | 1.4 | 1.5 | 1.3 |
| **Understanding** |  |  |  |  |  |  |  |  |  |  |  |  |  |  |
| I think this food labelling is: |  | **p= 0.0450** |  |  | **p= 0.2223** |  |  |  | **p= 0.0090** |  |  | **p= <0.0001** |  |  |
| Very easy/easy to understand | 73.9 | 72.7 | 74.9 | 79.1 | 78.3 | 75.8 | 75.7 | 75.5 | 66.5 | 74.1 | 77.8 | 69.1 | 77.0 | 82.3 |
| Neither hard nor easy to understand | 17.4 | 20.5 | 17.5 | 12.2 | 11.4 | 14.2 | 16.5 | 18.6 | 22.2 | 16.7 | 15.2 | 20.8 | 16.6 | 10.8 |
| Hard/very hard to understand | 8.7 | 6.8 | 7.6 | 8.7 | 10.4 | 10.1 | 7.8 | 5.9 | 11.3 | 9.2 | 6.9 | 10.1 | 6.3 | 6.9 |
| One food has 3 warnings, another has 1, which is healthier? |  | **p= 0.05** |  |  | **p= 0.23** |  |  |  | **p= 0.001** |  |  | **p= <0.0001** |  |  |
| The one with 3 warnings is healthier | 4.6 | 5.1 | 2.9 | 1.9 | 3.0 | 2.8 | 3.7 | 2.4 | 6.5 | 2.7 | 2.7 | 5.7 | 2.0 | 2.0 |
| Both foods are equally healthy | 9.4 | 7.7 | 8.0 | 9.8 | 7.9 | 10.4 | 10.1 | 6.3 | 13.3 | 7.6 | 8.3 | 12.5 | 7.4 | 6.0 |
| The one with 1 warning is healthier | 86.0 | 87.2 | 89.1 | 88.4 | 89.0 | 86.8 | 86.2 | 91.3 | 80.2 | 89.7 | 89.0 | 81.8 | 90.1 | 92.0 |
| Have you seen or heard on the media information about the FOPWL? |  | **p= 0.3769** |  |  | **p= 0.03** |  |  |  | **p= 0.02** |  |  | **p= 0.9490** |  |  |
| Yes | 51.0 | 53.7 | 49.3 | 50.6 | 42.2 | 50.2 | 55.1 | 49.8 | 41.4 | 50.6 | 52.7 | 51.3 | 51.5 | 50.6 |
| No | 49.0 | 46.3 | 50.7 | 49.4 | 57.8 | 49.8 | 44.9 | 50.2 | 58.6 | 49.4 | 47.3 | 48.7 | 48.5 | 49.4 |
| **Use** |  |  |  |  |  |  |  |  |  |  |  |  |  |  |
| How often have you compared the number of warnings between similar products? |  | **p= 0.3208** |  |  | **p= 0.5377** |  |  |  | **p= <0.0001** |  |  | **p= 0.0002** |  |  |
| Very often/Frequently | 23.1 | 18.7 | 21.5 | 24.8 | 18.3 | 20.3 | 21.9 | 23.6 | 12.9 | 16.0 | 25.4 | 19.4 | 20.2 | 27.3 |
| Sometimes | 42.2 | 46.7 | 43.2 | 40.1 | 42.6 | 43.3 | 45.7 | 40.6 | 40.1 | 44.6 | 42.5 | 40.1 | 46.5 | 42.3 |
| Rarely/Never | 34.7 | 34.6 | 35.3 | 35.1 | 39.2 | 36.4 | 32.5 | 35.8 | 47.0 | 39.4 | 32.1 | 40.5 | 33.3 | 30.4 |
| **Perceived impact** |  |  |  |  |  |  |  |  |  |  |  |  |  |  |
| What have you done if a product you used to buy for your child has warnings? |  | **p= 0.71** |  |  | **p= 0.37** |  |  |  | **p= 0.04** |  |  | **p= <0.0001** |  |  |
| Have continued to buy it as before | 8.6 | 6.8 | 7.4 | 9.9 | 5.4 | 8.5 | 7.7 | 8.8 | 12.1 | 8.2 | 7.5 | 9.8 | 8.9 | 5.3 |
| Have bought less than before | 63.4 | 64.4 | 66.7 | 63.5 | 59.1 | 66.7 | 64.3 | 65.1 | 68.2 | 67.2 | 63.4 | 62.6 | 68.2 | 61.7 |
| Have stopped buying it | 24.8 | 26.2 | 23.3 | 24.2 | 30.8 | 22.6 | 25.4 | 23.9 | 17.2 | 21.9 | 26.6 | 24.2 | 20.0 | 31.7 |
| Have not seen warnings on the products I buy | 3.2 | 2.6 | 2.5 | 2.4 | 4.7 | 2.2 | 2.6 | 2.3 | 2.5 | 2.7 | 2.5 | 3.5 | 2.9 | 1.3 |
| Have any of the warnings influenced your purchase decision more? |  | **p= 0.23** |  |  | **p= 0.05** |  |  |  | **p= <0.0001** |  |  | **p= 0.01** |  |  |
| Excess calories | 7.5 | 7.3 | 6.7 | 6.1 | 4.2 | 7.1 | 6.3 | 7.3 | 9.1 | 7.6 | 6.1 | 8.0 | 6.7 | 5.1 |
| Excess sodium | 6.8 | 7.5 | 6.7 | 6.2 | 4.5 | 4.5 | 8.5 | 7.8 | 7.6 | 5.6 | 7.0 | 6.4 | 6.8 | 7.0 |
| Excess trans fat | 7.1 | 4.9 | 5.5 | 9.6 | 5.1 | 5.7 | 7.9 | 6.3 | 9.1 | 8.2 | 5.8 | 5.6 | 5.8 | 9.4 |
| Excess sugars | 28.7 | 33.7 | 29.2 | 26.9 | 40.0 | 30.9 | 29.1 | 27.2 | 31.1 | 33.5 | 28.1 | 30.5 | 29.1 | 29.1 |
| Excess saturated fat | 9.2 | 9.3 | 8.7 | 8.7 | 6.1 | 12.8 | 7.3 | 8.8 | 5.9 | 11.9 | 8.5 | 9.1 | 9.8 | 7.5 |
| None | 9.5 | 7.4 | 10.8 | 9.0 | 8.1 | 8.1 | 8.2 | 10.8 | 13.4 | 8.7 | 8.6 | 11.0 | 10.0 | 6.0 |
| All equal | 31.2 | 29.9 | 32.3 | 33.5 | 32.0 | 30.9 | 32.8 | 31.7 | 23.7 | 24.6 | 36.0 | 29.4 | 31.8 | 35.9 |

\*adjusted by socioeconomic status, geographic region, parents' education level, nutrition consciousness, nutrition knowledge, BMI, age, sex, children's age, and sex.

### Understanding

Fifty-eight percent of parents strongly agreed/agreed that the FOPWL is easily understood by their children, with an increased understanding as children aged (49% among 5-6 y old, 60% among 7-8 y old, and 68% among 9-10 y old) (Figure 2). About their own understanding, most think that the FOPWL is very easy/easy to understand (74%). The proportion reporting that it is very easy/easy to understand was higher among those with high nutrition consciousness (78%) and high nutrition knowledge (82%) compared to their counterparts. The majority (86%) answered correctly that a product with one warning is healthier than a product with three warnings. This proportion was higher among those with high vs low nutrition consciousness (89 vs 80%) and high vs low nutrition knowledge (92 vs 82%). Finally, half of the sample have not seen or heard information about the FOPWL in the media, and this was higher among those with low vs high SES (58 vs 50%) and low vs high nutrition consciousness (59 vs 47%) (Table 2).

**Figure 2.**
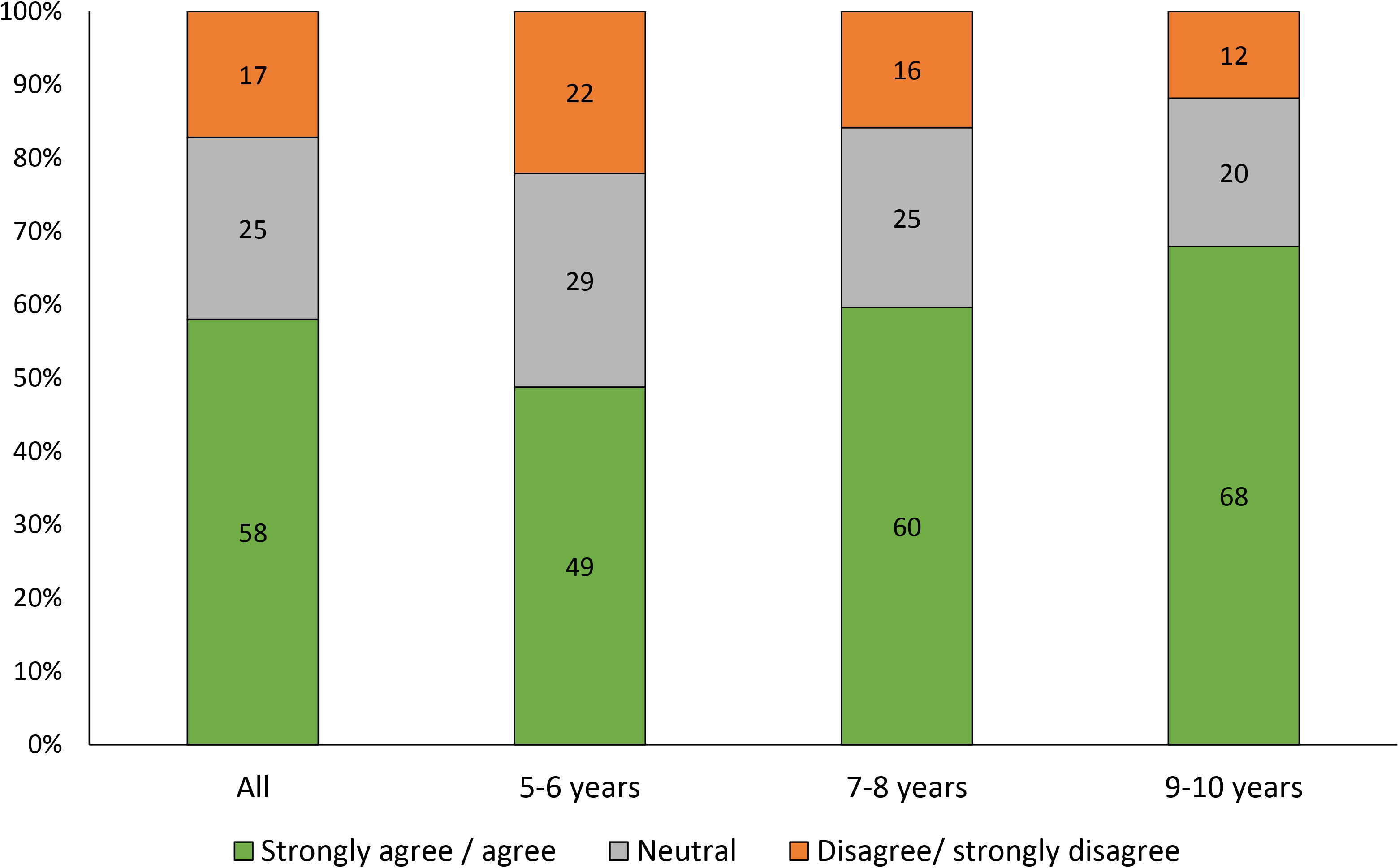
Parents’ level of agreement with the statement “my child understands easily the warnings labels” among all and by children’s age.* *adjusted by socioeconomic status, geographic region, parents’ education level, nutrition consciousness, nutrition knowledge, BMI, age, sex, and children’s sex.

### Use

To the question of how often parents compare the number of warnings between similar products, the answers were: 23% very often/frequently, 42% sometimes, 35% rarely/never. The proportion that compared very often/frequently the number of warnings was higher among those with higher compared to lower nutrition consciousness (25 vs 13%), and among those with higher compared to lower nutrition knowledge (27 vs 19%) (Table 2).

### Perceived impact

Most parents reported that if a product they used to buy for their child had warnings they have bought less (63%) or have stopped buying (25%), whereas 9% reported they have continued to buy it as before (Table 2 and Figure 3). The proportion of parents who reported stopped buying a product for their children was higher among those with higher compared to lower nutrition consciousness (27 vs 17%), and among those with higher compared to lower nutrition knowledge (32 vs 24%). When the question referred to a product they used to purchase for themselves, the proportion reporting they have continued to buy it as before was of 14% (Figure 3).

**Figure 3.**
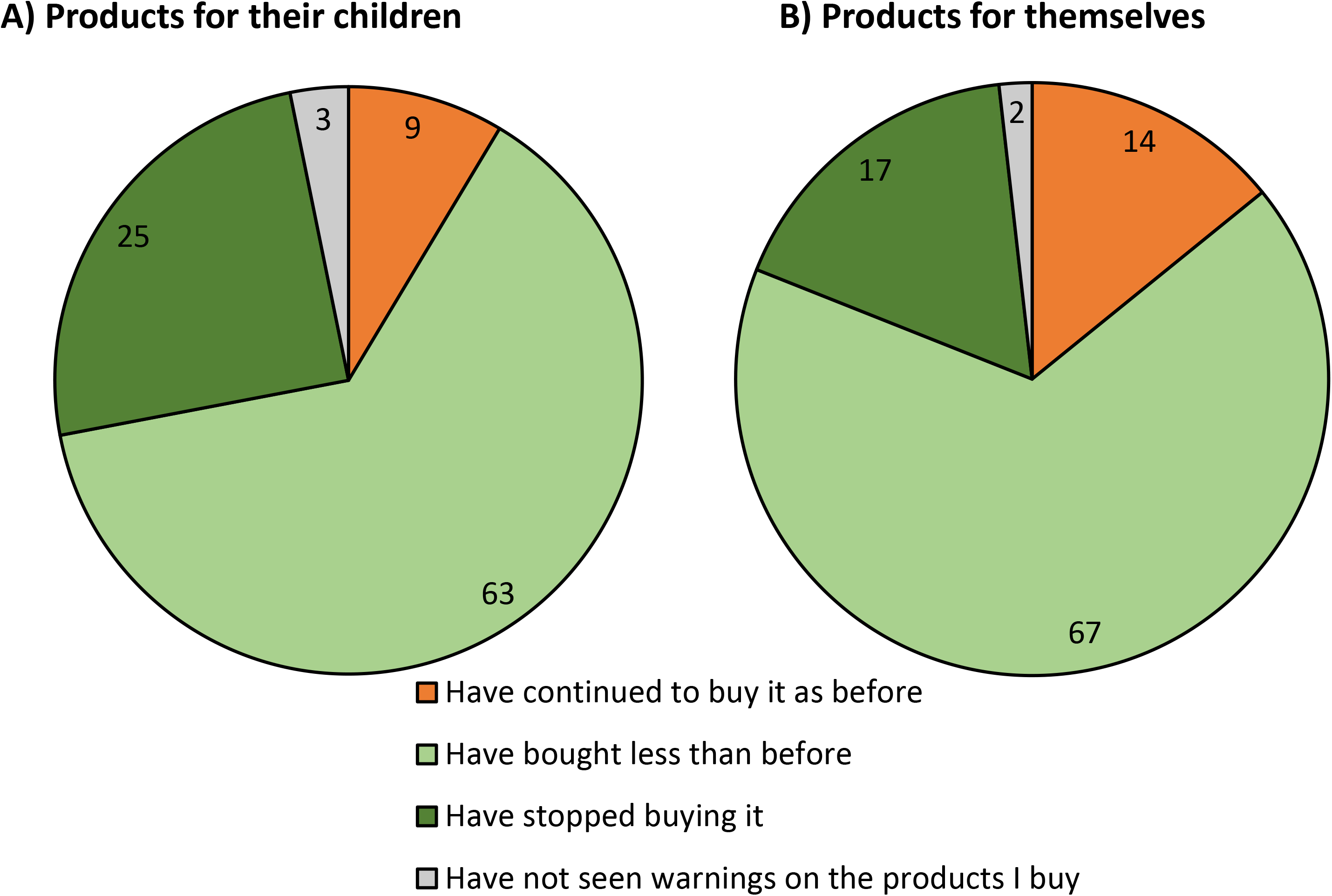
Perceived impact of FOPWL on purchases of a product with warnings parents used to buy A) for their children and B) for themselves.* *adjusted by socioeconomic status, geographic region, parents’ education level, nutrition consciousness, nutrition knowledge, BMI, age, sex, children’s age, and sex.

In total, 31% of parents reported that all nutrient warnings had influenced their purchasing decisions equally, whereas 29% reported that the “excess sugars” warning had influenced them more, and close to 10% that none of the warnings had influenced them (Table 2). Reported influence in the parent’s purchasing decision for the remaining warnings (calories, sodium, trans fat, or saturated fat) fluctuated from 6—9%. Among parents with low compared to high SES, the proportion reporting having been more influenced by the “excess sugar” was higher (40 vs 27%), while the proportion influenced by the other “excess-in” nutrients was lower (4—6 vs 6—9%).

Among those with higher nutrition consciousness or knowledge, the proportion reporting that all warning labels influenced them equally was higher (36%) compared to those with lower nutrition consciousness or knowledge (24 and 29%, respectively).

Before the FOPWL was implemented, more than half of the participants used to purchase sodas, breakfast cereals, and yogurt; <20% used to purchase cereal bars; and 20-50% used to purchase the remaining categories (Figure 4). Among those who used to purchase each food category, ∼70% (across categories) reported changing their purchases because of the FOPWL (∼50% purchased less, ∼12% switched to products with fewer warnings, ∼7% stopped purchasing it). Within food categories, sodas, juices and cereal bars had a higher proportion of participants changing their purchases (∼75%). Cereal bars, in particular, had the highest proportion of participants reporting having stopped purchasing them (16%). For the remaining food categories, <70% reported changing their purchasing decisions. Chips and chocolate powder were the food categories in which a higher proportion reported not changing their purchases because it was their favorite brand; whereas peanuts, yogurt, cheese, processed meat, and corn “tostadas” were the food categories in which a higher proportion reported not changing because they thought these had few/none warnings.

**Figure 4.**
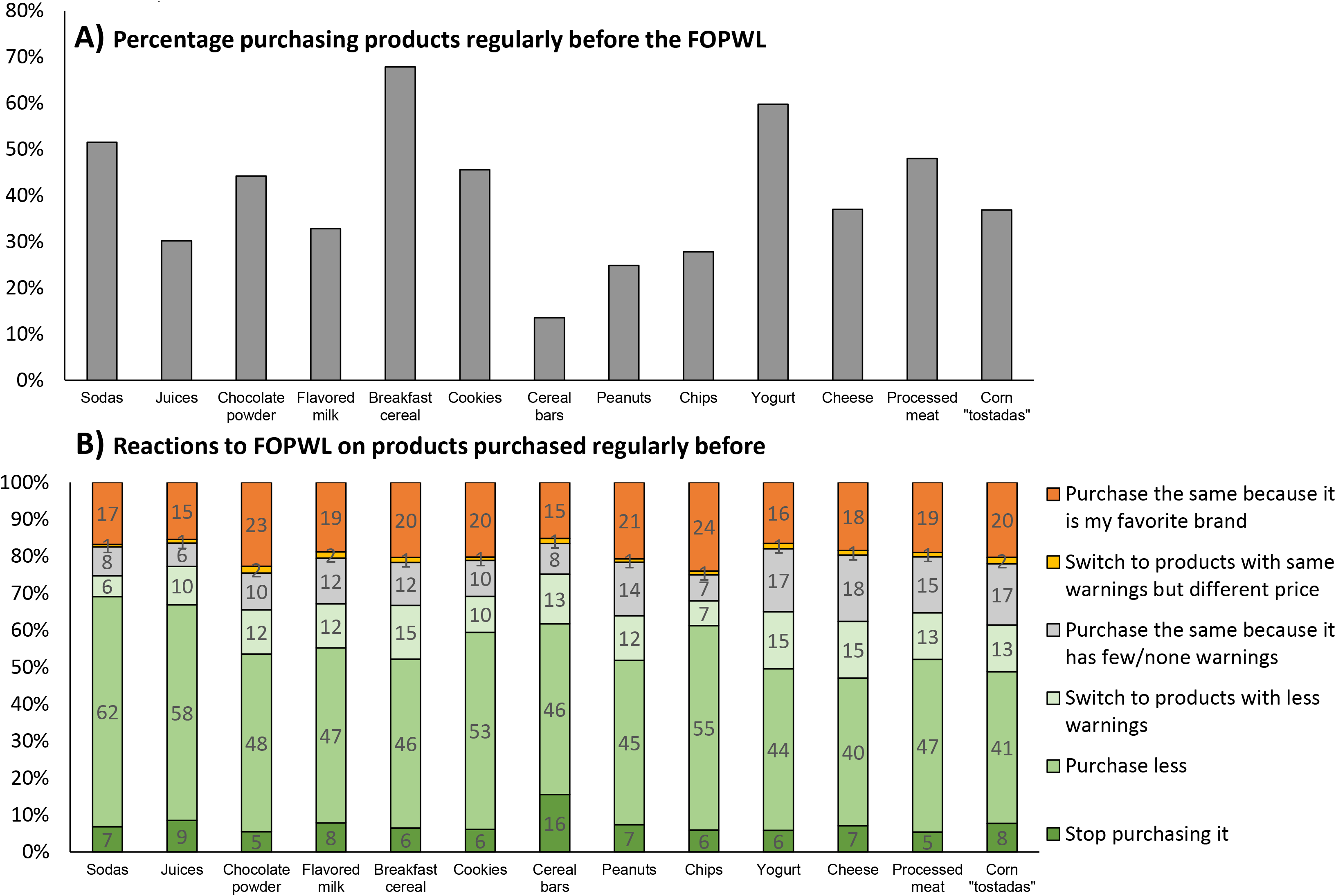
Perceived impact of FOPWL on purchases of products parents used to buy regularly before by food category.* *adjusted by socioeconomic status, geographic region, parents’ education level, nutrition consciousness, nutrition knowledge, BMI, age, sex, children’s age, and sex.

## DISCUSSION

In this study among Mexican parents of school-aged children, conducted within six months of the labelling law’s implementation, we found favorable reactions to the FOPWL. Results showed that around 85% of parents supported the FOPWL, 86% correctly understood it (i.e., objective understanding), 65% used it at the time of purchasing food products, and 88% perceived the warnings had influenced their buying decisions. The perceived impact was higher when products were for their children than for themselves, and higher for sodas, juices, and cereal bars and lower for chips and chocolate powder. We found that the responses were more favorable for most questions among those with higher nutrition knowledge or nutrition consciousness, and for few of the questions among those with higher education level.

Our findings are similar to those of a survey conducted in Chile six months after the FOPWL implementation [26]. The survey in Chile was a probabilistic sample of 1,067 adults representative of the general population, thus it was not restricted to parents of school-aged children or subjects from middle and high SES as in our study. Yet, in line with our data indicating high support for FOPWL among Mexican parents, most Chilean participants (92%) evaluated the FOPWL as good or very good. In Chile, among those that indeed used or were influenced by the FOPWL, ∼70% perceived that they chose products with fewer warnings, ∼15% bought less, and ∼10% did not buy products with warnings. In contrast, among this Mexican sample, those that were influenced by the FOPWL, only ∼15% changed to products with fewer warnings, the majority (∼70%) bought less, and as in Chile, ∼10% stopped purchasing products with warnings. Hence, in Chile the prevailing change was to choose products with fewer warnings, whereas in Mexico it was to buy less. Possibly, given the stricter nutrient cutoff points used in Mexico, there was less variation in the number of warnings within food categories in Mexico compared to Chile [31]. Also, the Ministry of Health from Chile launched a communication campaign that prompted consumers to choose products with fewer warnings [28]. Furthermore, the influence of each specific warning label was very comparable between Chile and Mexico. In both countries around 30-40% were influenced equally by all warnings, 20-30% were more influenced by the sugar warning label, around 10% were more influenced by one of the remaining labels, and 10% were not influenced by any of the nutrient warnings.

Interestingly, in Chile those with higher SES were more influenced by the sugar warning, while in Mexico this happened among those with lower SES. Finally, in Chile households with children reported comparing foods using the FOPWL during their purchases more frequently compared to households without children (47 vs. 40%). Similarly, in Mexico, the perceived impact of parents was higher if the product was for their children rather than for themselves.

By food category we found that parents perceived a higher impact in their purchases for sodas, juices, and cereal bars. Only for juices these results are in line with Chile’s, as the highest decrease in purchases was for fruit drinks, followed by dairy and lastly by sodas [27]. As suggested by the expectancy disconfirmation theory, the largest impact of FOPWL is generally observed for food categories in which the FOPWL gives the newest and most unexpected information regarding their nutritious characteristics [12,28]. However, FOPWL might also work as a reminder of the health harms of products already perceived as unhealthy [12]. In our findings, for juices and cereal bars it is likely that expectancy disconfirmation was the driver of the large perceived impact, as these are commonly misperceived as healthy products by the population, whereas for sodas it could be more related to the reminder mechanism. For instance, in a previous experimental study in Mexican population, the FOPWL made the lowest difference in correctly identifying unhealthy items for sugary drinks, suggesting that the FOPWL was giving the least new information for these beverages[14].

Socioeconomic characteristics and previous knowledge or attitudes in nutrition are all important factors than can modify the effect of FOPWL on purchases behavior, as described in the conceptual framework proposed by Smith Taillie et. al.[7]. Given that the FOPWL works through increasing awareness of the nutrient composition of foods and is a form of educating the consumer, it is expected that this type of policy will be taken up more easily by those who already have interest in the topic. In contrast, other policies such as taxes or school/public spaces food access regulations would be less influenced by the baseline motivation or resources of individuals [21]. In a previous randomized experiment in Mexican population, those with higher income, education, and self-perceived nutrition knowledge made healthier food choices when exposed to the FOPWL compared to their respective counterparts [15]. In our study, the influence of nutrition knowledge or consciousness on having favorable responses to the FOPWL was much larger than that of education or SES. By education, only in few questions were more favorable among those with higher education. By SES the differences were not in a clear direction, those with lower SES were less neutral in their support to the FOPWL (they agree and disagree more with the FOPWL), they were more influenced by the excess sugar warning, and a higher proportion had not seen information about the FOPWL in the media. These small differences by education or mixed results by SES should be confirmed in a sample including a wider range of SES stratums, but previous evidence in Mexico suggests that the FOPWL is the most effective across SES stratums in comparison to other labelling systems [13–15]. In addition, the more favorable response to the FOPWL by those with higher nutrition education or consciousness could have been exaggerated due to a higher social desirability bias among this stratum. Nevertheless, nutrition knowledge and consciousness might play an important role, and efforts to enhance these in the population are needed. For instance, there is a new required course named “Healthy life” in the Mexican basic education system which aims to educate and promote healthy lifestyles, including dietary habits.

The FOPWL until now has been implemented without a complementary national education campaign and was also not accompanied with the strengthening of other policies like marketing or school regulations as in Chile. Yet, there was a massive campaign launched by Civil Society Organizations and a lot of interest on FOPWL by the media. We found that about half of our sample heard information of any form about the new FOPWL. It would be of interest to identify the effect of a national educational campaign released years after implementation. On the one hand the window of opportunity when the FOPWL is a novelty has passed, but on the other hand some time is needed to correct all errors in the FOPWL and sanction uncompliant companies; a campaign released when errors have not been revised could lead to confusion. From a research stand point, having variation in the specifics of how FOPWLs are implemented across countries could help us identify the most effective implementation strategies and the role of each strategy.

Our study assessed the self-reported reactions to the FOPWL of the participants of this cohort, which might reflect intention to change rather than actual change; it is not an impact evaluation of the FOPWL on behaviors or health indicators. Notwithstanding, the FOPWL was implemented during the COVID-19 pandemic, which also influenced population’s dietary habits [32,33]. According to an online survey carried out in Mexico during the first year of the pandemic, 42% of participants perceived that their intake of healthy foods increased, and 40% that their intake of unhealthy foods decreased [32]. At the same time, in this period of social distancing and confinement, the food industry had aggressive marketing campaigns [34]. Therefore, isolating the impact of the FOPWL from the COVID-19 pandemic might pose a challenge, so studies like ours gain importance by informing about the degree of acceptance to the FOPWL, a key intermediate outcome.

This study has several limitations and strengths. Our sample is not representative of the Mexican population, as participants were limited to middle and high SES. This was expected because both recruitment and data collection were conducted online. However, an important advantage of conducting the study online was that participants were distributed across the 32 states of the country. Self-selection is also a limitation; it is possible that parents interested in nutrition were more likely to join the study. We put in place several strategies to limit the selectivity of the sample: first, we did not recruit participants through our institutional social media accounts because the followers are typically nutrition professionals, general public highly interested in nutrition, or both advocates and opponents of food policies; second, the recruitment strategy in social media platforms was mainly through paid advertisements, which can reach a population of more varied backgrounds and lifestyles in comparison to reaching only the followers of specific accounts; and third, incentives (gift cards and electronic tablet’s raffles) are given for participation, which gives motives, other than nutrition/health related, for participation. Furthermore, by measuring the level of nutrition knowledge and consciousness, we were able to stratify and identify the response to the FOPWL across different stratums. Finally, overestimation of support, use and perceived impact, particularly driven by social desirability bias, is a potential issue.

In sum, in this national web-based cohort of parents of school-aged children we found that within 6 months of implementation, the immediate self-reported responses to the Mexican FOPWL were favorable. The perceived impact of the FOPWL was higher when products were for their children than for themselves, and higher for sodas, juices, and cereal bars categories and lower for chips and chocolate powder. There were some differences by individual’s characteristics; mainly those with higher nutrition knowledge or nutrition consciousness responded more favorably to the FOPWL. In fewer cases, those with higher education had more favorable responses to the FOPWL. Yet, the overall support, use, perceived impact, and understanding of the FOPWL was high across all segments of the studied population. More studies, on different outcomes, populations, and over longer terms, are required to obtain a fuller picture of the response to the FOPWL in Mexico.

## List of abbreviations

FOPWL: Front-of-package warning labelling
SES: Socioeconomic Status

## Declarations

### Ethics approval and consent to participate

Inform consent was obtained from each participant at the beginning of the survey. The survey protocol was reviewed and approved by the Research and Ethics Committees of the Mexican National Institute of Public Health.

### Availability of data and materials

The datasets used and/or analyzed during the current study are available from the corresponding author on reasonable request.

### Competing interests

The authors declare that they have no competing interests

### Funding

Bloomberg Philanthropies.

## Data Availability

All data produced in the present study are available upon reasonable request to the authors

## Acknowledgements

We would like to thank the participants of the EPHA Niñ@s cohort for all their time; Patricia I. Meza Gordillo, MSc and Úrsula Monserrat Martínez Garza, MSc, Ph.D. for their support with participants recruitment; Omar Sainz Mendez for his work as field coordination; Oralia Villanueva Reyes, Luis H García Uribe, Jessica L. Alvarez Barrientos, Daniel Benítez Sánchez, María del Rocio Pérez Reyes, Angélica Flores Reyes and Valeria Cruz Villalba for data collection; Ángel A. García Avilés for programming the survey and helping with data managing, and Diana Lozano for administrative support.

## Notes

### Competing Interest Statement

The authors have declared no competing interest.

### Funding Statement

This study was funded by Bloomberg Philanthropies.

### Author Declarations

The survey protocol was reviewed and approved by the Research and Ethics Committees of the Mexican National Institute of Public Health.

